# Rare Variants in Antisense lncRNA-Protein Coding Gene Overlap Regions Contribute to Obsessive-Compulsive Disorder

**DOI:** 10.1101/2025.03.07.25323592

**Authors:** Seulgi Jung, Madison Caballero, Shelby Smout, Behrang Mahjani

## Abstract

Obsessive-compulsive disorder (OCD) is a prevalent neuropsychiatric disorder with an incompletely understood genetic basis, limiting targeted therapeutic options. Although previous rare variant studies have primarily focused on protein-coding genes, the contribution of rare regulatory non-coding variants remains largely unexplored. We analyzed whole-genome sequencing data from 2,561 OCD cases and 12,974 controls from the All of Us Research Program to investigate rare, conserved variants (minor allele count ≤ 5, GERP++ > 0) within antisense long non-coding RNA (lncRNA) and protein-coding gene overlap regions. A burden analysis identified a significant association with OCD in the *KNCN/MKNK1-AS1* overlap region (odds ratio: 5.1, FDR < 0.05). Expression analysis revealed strong co-expression of these genes in striatal brain regions implicated in OCD pathophysiology. Genes co-expressed with *KNCN/MKNK1-AS1* were enriched for synaptic vesicle dynamics, calcium signaling, and established OCD risk genes, highlighting the importance of non-coding regulatory variation in psychiatric genetics.

## Introduction

Obsessive-compulsive disorder (OCD), characterized by intrusive thoughts and repetitive behaviors, affects 1–3% of the global population and typically manifests in childhood. Current therapeutic options offer limited efficacy, partly due to an incomplete understanding of the disorder’s biological basis.^1–3^ Highly conserved neurodevelopmental processes and cortico-striatal-thalamo-cortical circuits are strongly implicated in OCD pathophysiology.^4,5^ However, while genome-wide association studies have identified several protein-coding risk loci, these explain only a fraction of OCD heritability, suggesting contributions from unexplored regulatory elements.^6–8^

Long non-coding RNAs (lncRNAs), transcripts longer than 200 nucleotides without protein-coding potential, have emerged as critical regulators of gene expression through mechanisms including chromatin remodeling, enhancer modulation, RNA processing, and nuclear architecture.^9,10^ Approximately 40% of identified lncRNAs display tissue-specific expression in the mammalian nervous system, underscoring their roles in neurodevelopment and brain function.^11,12^ Increasing evidence implicates dysregulated lncRNAs in various neuropsychiatric disorders, including autism spectrum disorder,^13^ Rett syndrome,^14^ schizophrenia,^15^ and fragile X syndrome^16^ highlighting the potential of these transcripts in shaping neurodevelopmental pathways.

A particularly neglected layer of regulation involves lncRNAs that overlap protein-coding genes in antisense orientation. These transcripts occupy a unique regulatory space: although they are non-coding, they overlap exons or introns of protein-coding genes, and a single nucleotide change can affect both transcripts.^17^ Antisense lncRNAs modulate the expression of proximal genes through a variety of mechanisms, including transcriptional interference, where antisense transcription impedes the progression of RNA polymerase on the sense strand, and epigenetic modifications, such as the recruitment of chromatin-modifying enzymes that alter histone modifications or DNA methylation patterns, thereby influencing gene expression.^18^ The regulatory specificity of antisense lncRNAs positions them as attractive therapeutic targets, amenable to interventions such as antisense oligonucleotides,^19^ small interfering RNAs,^20^ or CRISPR-based RNA editing,^21^ potentially offering enhanced specificity and reduced side effects compared to traditional pharmacotherapies.^3,22^

To address this gap, we analyzed whole-genome sequencing (WGS) data from 2,561 OCD cases and 12,974 controls in the All of Us Research Program.^23^ We hypothesized that rare conserved variants within antisense lncRNA-protein coding gene overlap regions contribute to OCD susceptibility. Specifically, we predicted that variants in these overlap regions would be enriched in OCD cases compared to controls, as such variants could simultaneously affect both antisense lncRNAs and protein coding genes, potentially producing larger functional effects than variants affecting either transcript alone. We further hypothesized that identified risk variants would occur in genes showing brain-specific expression patterns relevant to OCD pathophysiology. Our analysis revealed significant enrichment of rare conserved variants in the *KNCN/MKNK1-AS1* overlap region, with functional implications for OCD pathogenesis. These findings illuminate a novel regulatory mechanism in OCD and suggest new avenues for targeted therapeutic development in neuropsychiatric disorders.

## Results

Our primary goal was to characterize the landscape of rare conserved genetic variants within the overlapping region between antisense lncRNAs and protein-coding genes, and evaluate their association with OCD risk. We acquired WGS data from 2,561 OCD-affected individuals and 12,974 ancestry-matched controls from the All of Us Research Program (see Methods).

We systematically identified variants occurring within genomic intervals where antisense lncRNAs overlap protein-coding genes, as annotated in GENCODE v47.^11^ We defined rare variants as those with minor allele count ≤ 5 in both the case-control cohort and the non-psychiatric subset of the gnomAD database (v3.1),^24^ corresponding to a maximum allele frequency of 0.015%. Our analysis focused exclusively on non-deleterious coding variation by excluding all protein-truncating variants and missense variants affecting the overlapping protein-coding sequences.

We implemented stringent quality control filters to ensure high-confidence variant calls applying minimum sequencing depth of 10x, mapping quality score >20, and other variant quality metrics. To enrich for functionally relevant variants, we utilized the genomic evolutionary rate profiling (GERP++) score, which quantifies evolutionary constraint at genomic positions based on conservation across species, with higher scores indicating stronger conservation and potential functional importance.^25^

We restricted our analysis to evolutionary conserved positions, defined as having a GERP++ score > 0. This conservation filter retained 79,209 rare variants (30% of the initial 263,322 variants), effectively balancing sensitivity with specificity to maintain adequate statistical power. To evaluate how different conservation stringency thresholds might influence our findings, we conducted sensitivity analyses using stricter thresholds (GERP++ > 1, > 2, and > 3). While higher conservation thresholds significantly reduced variant numbers, retaining fewer than 10% of variants at GERP++ > 1 (Supplementary Table 1). These stricter thresholds did not substantially alter the overall relative risk estimates or the statistical significance of our primary findings, reinforcing our selection of GERP++ > 0 as the optimal threshold.

Across 992 antisense lncRNA–protein-coding gene overlap regions, we identified 13,091 rare variants in cases and 66,118 rare variants in controls. Initial burden analysis revealed no significant difference in the overall distribution of rare variants between cases and controls (odds ratio[OR] = 1.00, binomial *P* = 0.75).

We examined the distribution of rare variant types across antisense lncRNA regions, protein-coding gene regions, and their overlapping intervals to identify potential enrichment patterns in OCD cases versus controls (Figure 1 and Supplementary Table 2). The variant spectrum was dominated by intronic variants in all three genomic contexts, comprising 88% of variants in antisense lncRNAs, 92% in protein-coding genes, and 82% of intronic variants in both antisense lncRNAs and protein-coding genes. We observed no significant differences in the distribution of any variant type between cases and controls across all three genomic contexts (binomial *P* > 0.05).

**Figure 1.**
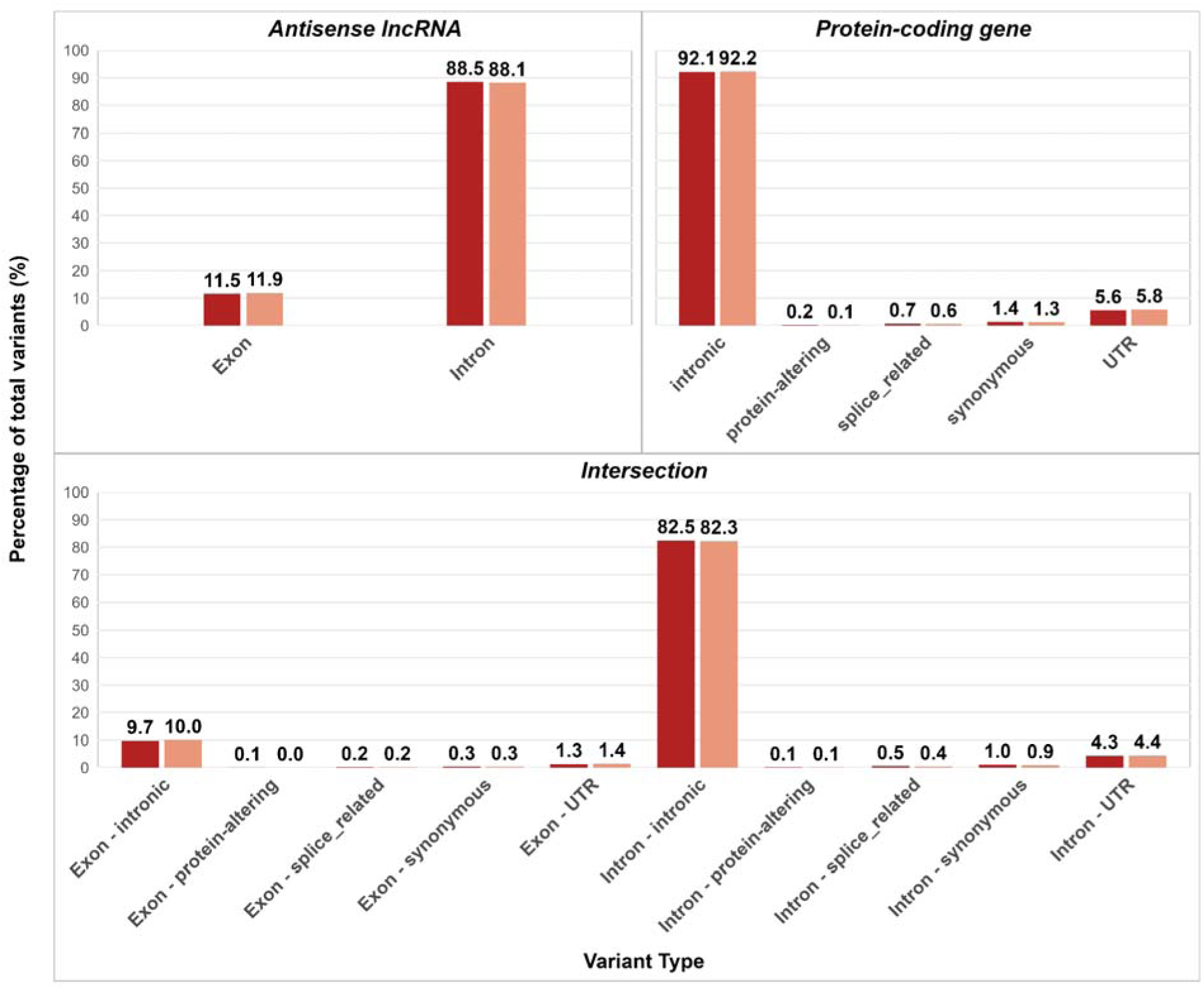
Distribution of rare variant types in antisense lncRNA and protein-coding gene regions in OCD cases and controls. Bar plots showing the percentage of rare variants by functional category in (A) antisense lncRNA regions, (B) protein-coding gene regions, and (C) overlapping regions between antisense lncRNAs and protein-coding genes. Dark blue bars represent OCD cases; light blue bars represent controls. Variant categories include exonic variants, intronic variants, and splice-related variants. Numbers above bars indicate the percentage of total variants in each category. No significant differences were observed between cases and controls for any variant type across all three genomic contexts (binomial *P* > 0.05). The analysis included 79,209 rare variants that passed quality control filters and showed evidence of evolutionary conservation (GERP++ > 0) across 992 antisense lncRNA-protein coding gene overlap regions.

To investigate whether the constraint level of protein-coding genes influences the burden of rare variants in antisense lncRNA overlap regions, we stratified genes into 20 quantiles based on their LOEUF (Loss-of-function Observed/Expected Upper bound Fraction) scores.^24^ LOEUF quantifies gene-level constraint, with lower scores indicating greater intolerance to loss-of-function variation, suggesting constrained genes. Notably, we observed a significant enrichment of rare conserved variants in OCD cases specifically within overlap regions of the least constrained genes (highest LOEUF quantile: relative risk = 1.35, 95% CI: 1.12-1.63, binomial *P* = 0.003; Figure 2).

**Figure 2.**
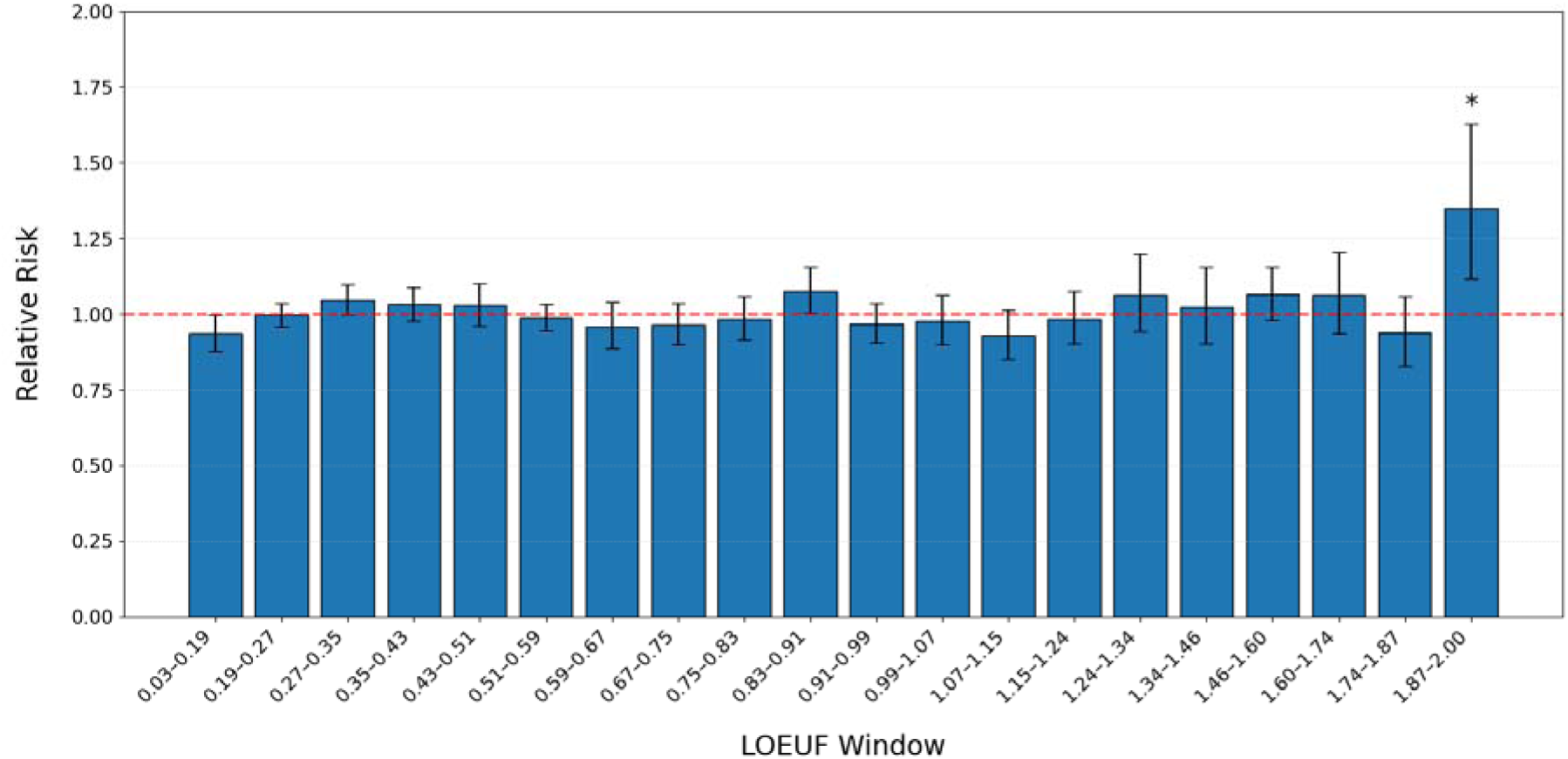
Enrichment of rare conserved rare variants within the overlapping region of antisense lncRNA and their sense protein-coding genes in OCD. Relative risk of rare conserved variants in the overlapping region across 20 equally-spaced quantiles of LOEUF scores. The first quantile includes most constrained genes. The figure shows risk ratios with 95% confidence intervals. The dashed horizontal line represents relative risk = 1, indicating no effect. Points above this line suggest increased risk. \**P*<0.05 using a binomial test between OCD cases and unaffected controls.

To examine whether the enrichment of rare variants in the least constrained genes was influenced by evolutionary conservation stringency (GERP++ score), we repeated our analysis using increasingly strict conservation thresholds (Supplementary Figure 1a-e). As we applied more stringent conservation filters—from including all variants regardless of the conservation score to requiring higher conservation scores (>0, >1, >2, and >3)— we observed a progressive increase in the relative risk for variants in the 20th LOEUF quantile.

This dose-dependent relationship between conservation score of rare variants and effect size in the least constrained genes highlights that highly conserved sequences within antisense lncRNA overlap regions may be particularly relevant for OCD risk. The enrichment specific to genes with high LOEUF scores suggests these less constrained protein-coding genes may be more sensitive to regulatory variation, possibly due to inherently more variable or context-dependent expression patterns.

### Antisense lncRNA and protein-coding gene pairs associated with the risk of OCD

To identify specific antisense lncRNA-protein coding gene overlap regions associated with OCD risk, we performed comprehensive rare variant association analyses using both Fisher’s exact and SKAT-O (sequence kernel association test-optimal) tests on 2,728 OCD cases and 13,517 controls to ensure robust detection across different genetic architectures. We analyzed cumulative rare variant burden across 992 overlapping regions between antisense lncRNAs and their corresponding protein-coding genes (Supplementary Table 3).

The quantile-quantile (QQ) plots revealed deflated genomic inflation factors in both analyses (Fisher’s exact test: λ_GC_ = 0.577; SKAT-O: λ_GC_ = 0.962), with values below the expected null of 1.0 (Figure 3a and b). This deflation likely reflects the sparsity of rare conserved variants within individual overlap regions and the focused nature of our analysis on specific genomic features (antisense lncRNA overlaps), where only a subset may be functionally relevant to OCD. The SKAT-O test showed less deflation than Fisher’s exact test, suggesting it may be more powerful for detecting associations when analyzing sparse variant data. Among all tested regions, a gene pair (*KNCN/MKNK1-AS1*) showed a significant association after multiple testing corrections using false discovery rate (FDR) < 0.05 (Table 1 and Figure 3a and b).

**Figure 3.**
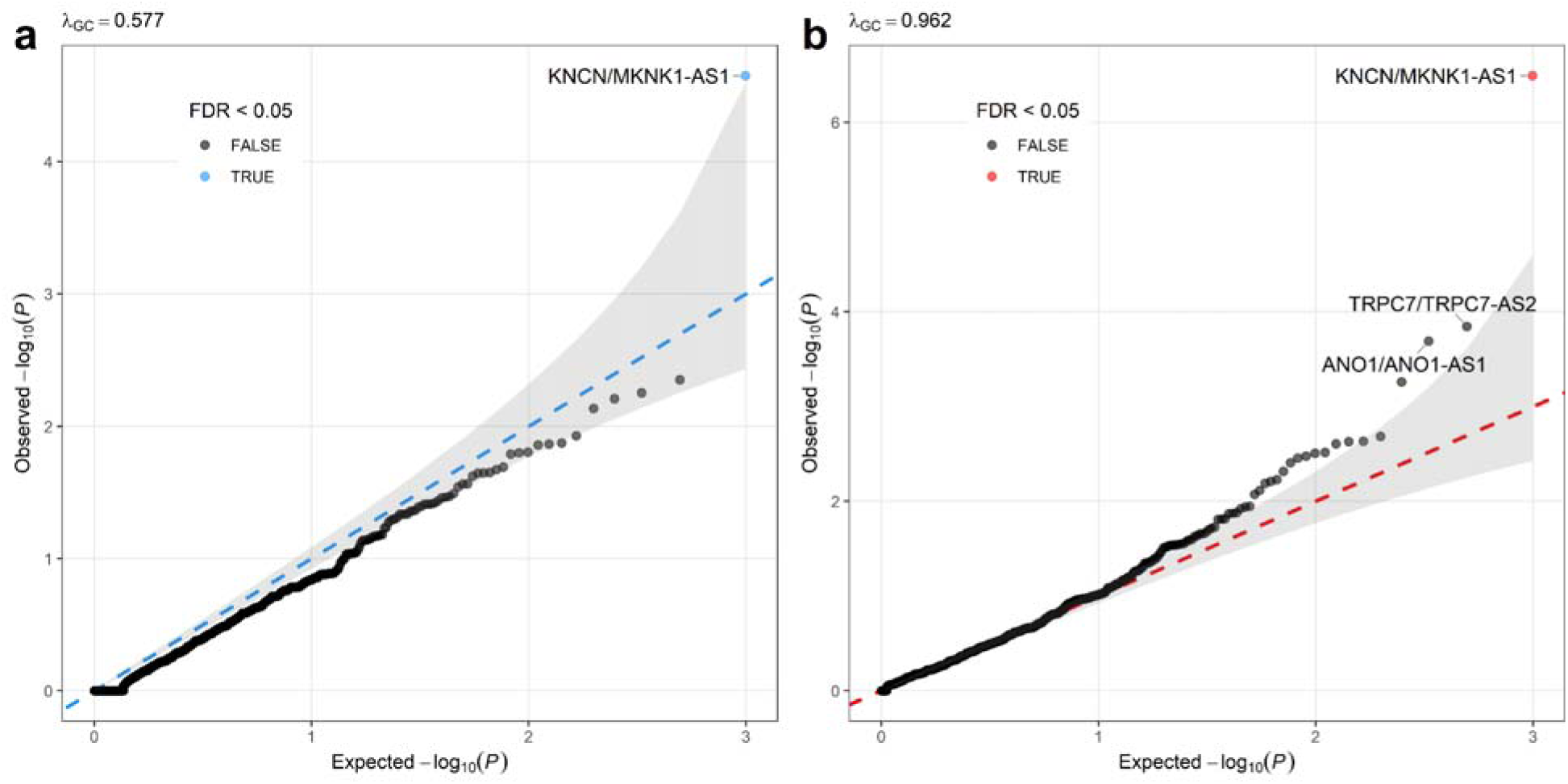
Quantile-quantile plots of rare variant association tests in antisense lncRNA-protein coding gene overlap regions. The quantile-quantile (QQ) plot displays the observed -log_10_(*P*) values against the expected -log_10_(*P*) values under the null hypothesis. The QQ plots show the distribution of association P-values for rare variant burden analysis in antisense lncRNA-protein coding gene overlap regions. (a) Fisher’s exact test results across 992 overlap regions (λ_GC_= 0.577). (b) SKAT-O test results for the same regions (λ_GC_ = 0.962). In both panels, the diagonal dashed line represents the null expectation of no association, with the gray shaded area indicating the 95% confidence interval. Points are colored based on false discovery rate (FDR) significance: blue/red points indicate FDR < 0.05, while gray points indicate FDR ≥ 0.05. The names of gene pairs with FDR < 0.1 are labeled.

**Table 1.**
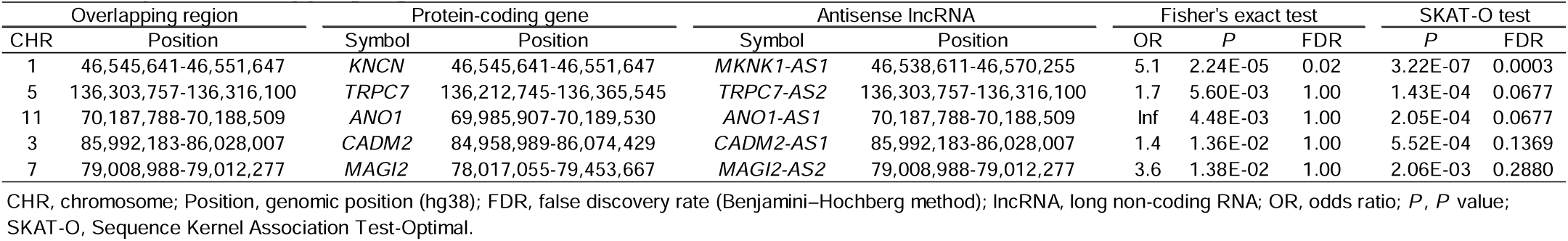
Top five overlapping regions in Fisher’s exact and SKAT-O test between 2,561 OCD cases and 12,974 controls.

*KNCN* encodes Kinocilin, highly expressed in brain tissues, while *MKNK1-AS1* (MKNK1 Antisense RNA 1) is an antisense long non-coding RNA that overlapped with both *KNCN* and *MKNK1* (MAPK Interacting Serine/Threonine Kinase 1). Rare variants associated with OCD were located at the 5′ end, 3′ end, and intronic sequences of *KNCN*, as well as intronic regions of *MKNK1-AS1* (Supplementary Figure 2).

Notably, rare conserved variants in the *KNCN/MKNK1-AS1* overlap region are within candidate cis-regulatory elements identified by ENCODE as having a high likelihood of being involved in regulating gene expression,^26^ transcription binding sites identified by JASPAR,^27^ and regulatory region identified by open regulatory annotation (ORgeAnno) database (Supplementary Figure 2).^28^ Single-nucleotide polymorphisms (SNPs) in this overlapping region showed expression quantitative trait loci (eQTL) effects on *KNCN*, *MKNK1-AS1*, *MKNK1*, *MOB3C*, *TMEM275*, *FOXD2*, and *EFCAB14* in brain and whole blood tissues (Supplementary Table 4).

Additionally, two overlap regions approached nominal significance (FDR < 0.1 in SKAT-O test) and warrant further investigation as candidate risk loci (Table 1 and Figure 3b). The *TRPC7/TRPC7-AS2* region (FDR = 0.0677 in SKAT-O) contains *TRPC7* (transient receptor potential cation channel subfamily C member 7), which encodes a calcium-permeable cation channel crucial for neuronal excitability and implicated in seizure susceptibility and circadian rhythm regulation.^29^ The *ANO1/ANO1-AS1* region (FDR = 0.0677 in SKAT-O) harbors *ANO1* (anoctamin 1), encoding a calcium-activated chloride channel essential for neuronal signaling.^30^ The near-significant associations of these functionally relevant genes suggest they may contribute to OCD pathophysiology through disrupted ion channel function and neuronal excitability.

### Gene expression correlation analysis

To investigate the potential regulatory relationship between the antisense lncRNA *MKNK1-AS1* and its overlapping protein-coding gene *KNCN*, we performed correlation analysis of their expression patterns across 13 brain regions from the GTEx database version 10 (Figure 4a and Supplementary Figure 3). We observed substantial variation in the strength of co-expression between these genes across different brain tissues.

**Figure 4.**
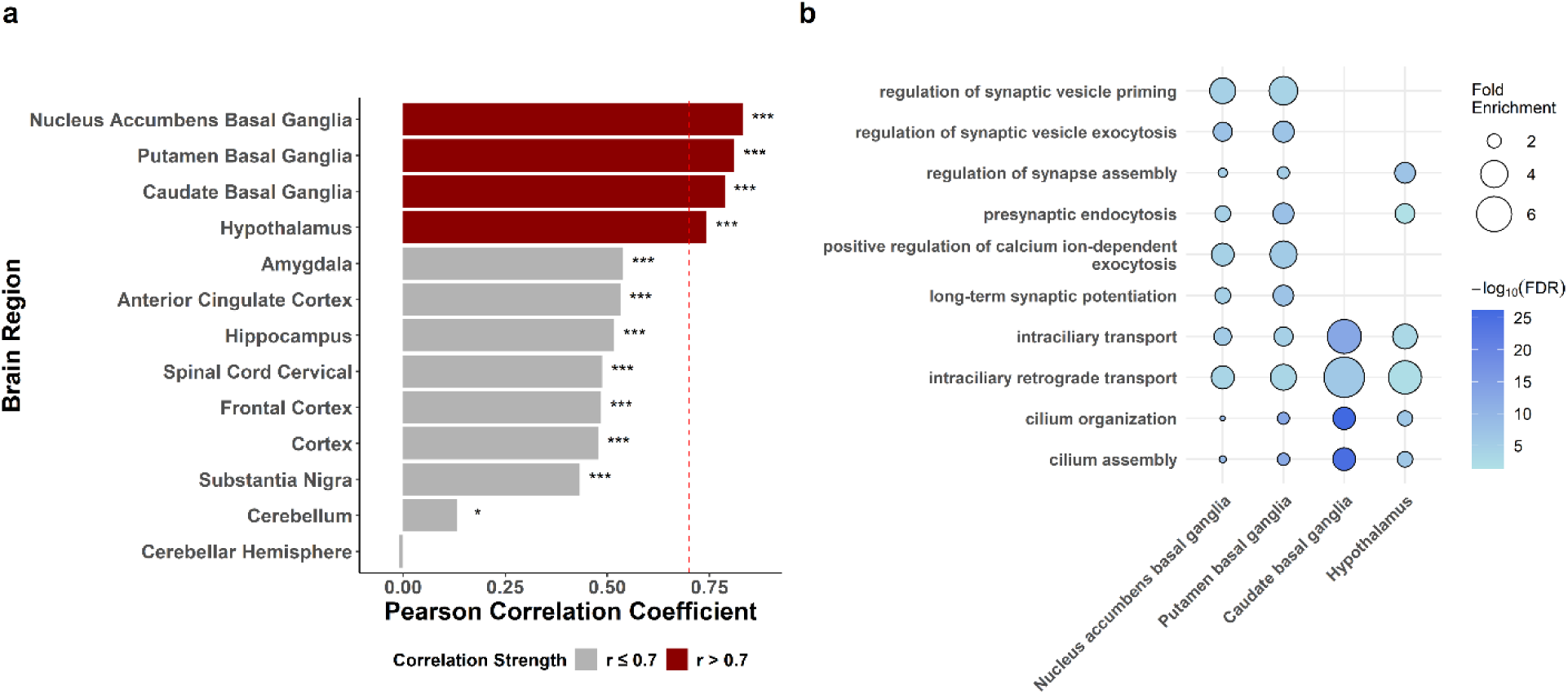
Brain region-specific co-expression analysis of *KNCN* and *MKNK1-AS1* and functional pathway enrichment. (a) Pearson correlation coefficients between *KNCN* and *MKNK1-AS1* expression levels across 13 brain regions from the GTEx v10 dataset. Brain regions are ordered by correlation strength. Dark red bars indicate strong positive correlations (r > 0.7), while gray bars indicate moderate to weak correlations (r ≤ 0.7). The dashed red line marks the r = 0.7 threshold. Significance levels are denoted as \**P*<0.05, \*\*\**P*<0.001. (b) Gene Ontology (GO) enrichment analysis of biological processes for genes showing strong positive correlation (r > 0.7) with both *KNCN* and *MKNK1-AS1* in the four brain regions with highest co-expression (nucleus accumbens, putamen, caudate, and hypothalamus). Circle size represents fold enrichment, and color intensity indicates -log_10_(FDR) values, with darker blue representing more significant enrichment.

A total of 12 brain regions showed positive correlation with *P* < 0.05 (Figure 4a and Supplementary Figure 3). The strongest positive correlations (r > 0.7) were observed in four brain regions critical for OCD pathophysiology: the nucleus accumbens (r = 0.83), putamen (r = 0.81), caudate (r = 0.79), and hypothalamus (r = 0.74). Notably, all three components of the striatum (the nucleus accumbens, putamen, and caudate) demonstrated remarkably high co-expression between *KNCN* and *MKNK1-AS1*, suggesting coordinated regulation within the basal ganglia circuitry.

To identify biological processes potentially influenced by the *KNCN/MKNK1-AS1* regulatory axis, we performed Gene Ontology (GO) enrichment analysis using genes that showed strong positive correlation (r > 0.7) with both *KNCN* and *MKNK1-AS1* expression in the four brain regions with highest co-expression (Supplementary Table 5). This analysis revealed significant enrichment for synaptic and neuronal function pathways, particularly those related to synaptic vesicle dynamics (regulation of synaptic vesicle priming; regulation of synaptic vesicle exocytosis), synaptic plasticity (long-term synaptic potentiation), and calcium signaling (positive regulation of calcium ion-dependent exocytosis) (Figure 4b and Supplementary Table 6). Additionally, we observed enrichment for cilium-related processes (cilium organization and cilium assembly) and intracellular transport mechanisms (intraciliary transport and intraciliary retrograde transport), suggesting potential roles in primary cilia function and cellular trafficking.

### Enrichment analysis of OCD risk genes

To determine whether OCD risk genes were enriched among genes co-expressed with *KNCN* and *MKNK1-AS1*, we used the 207 genes identified in the current OCD genome-wide association study (GWAS)^6^ as OCD risk genes and performed Fisher’s exact tests comparing their proportion within high-correlation gene sets versus tissue-specific background genes. All four brain regions showed significant enrichment of OCD risk genes with FDR < 0.05 among genes highly correlated with both *KNCN* and *MKNK1-AS1* expressions (Table 2). The nucleus accumbens demonstrated the most significant enrichment, followed by the caudate, hypothalamus, and putamen.

**Table 2.** Enrichment of OCD risk genes among genes highly correlated with both KNCN and MKNK1-AS1 expression.

| Tissue | r > 0.7 with both KCNC and MKNK1-AS1 |  | r < 0.7 with either KCNC or MKNK1-AS1 |  | *P* | OR | FDR |
| --- | --- | --- | --- | --- | --- | --- | --- |
|  | OCD risk genes | Non-OCD risk genes | OCD risk genes | Non-OCD risk genes |  |  |  |
| Nucleus accumbens basal ganglia | 57 | 6,287 | 84 | 18,709 | 5.21E-05 | 2.02 | 0.0002 |
| Caudate basal ganglia | 25 | 2,340 | 115 | 22,214 | 1.67E-03 | 2.06 | 0.0028 |
| Hypothalamus | 22 | 1,995 | 121 | 23,182 | 2.14E-03 | 2.11 | 0.0028 |
| Putamen basal ganglia | 44 | 5,043 | 94 | 18,532 | 2.77E-03 | 1.72 | 0.0028 |

FDR, false discovery rate; OCD, obsessive-compulsive disorder; OR, odds ratio; *P*, *P* value of Fisher's exact test

## Discussion

This study provides systematic evidence linking rare conserved variants in antisense lncRNA-protein coding gene overlap regions to OCD risk. Our findings reveal a genetic architecture where single nucleotide variants can simultaneously affect both regulatory lncRNAs and their overlapping protein-coding genes, potentially amplifying their functional impact on disease susceptibility.

The identification of *KNCN/MKNK1-AS1* as the top association signal is particularly intriguing given the biological functions of these genes. *KNCN* encodes kinocilin, a protein involved in stabilizing dense microtubular networks or in vesicular trafficking,^31^ while MKNK1 encodes a kinase that regulates translation in response to synaptic activity.^32^ The antisense lncRNA *MKNK1-AS1* overlaps both genes, creating a complex regulatory unit where variants could disrupt multiple aspects of neuronal function simultaneously. The localization of OCD-associated variants to conserved regulatory elements within this overlap region, coupled with their eQTL effects on multiple genes, suggests these variants may alter the coordinated expression of genes critical for neural circuit function.

Our observation that rare variant enrichment was specific to overlapping regions of less constrained genes (high LOEUF scores) provides important insights into the genetic architecture of OCD. This pattern suggests that while highly constrained genes are intolerant to variation due to their essential functions, less constrained genes may rely more heavily on precise regulatory control, making their antisense lncRNA regulatory mechanisms particularly vulnerable to disruption. The dose-dependent relationship between variant conservation and effect size further supports this model, indicating that highly conserved regulatory sequences within these overlap regions are critical for maintaining proper gene expression patterns.

The brain region-specific expression analysis revealed remarkably high co-expression between *KNCN* and *MKNK1-AS1* in striatal regions (nucleus accumbens, putamen, and caudate) and hypothalamus, all key components of the cortico-striato-thalamo-cortical circuits implicated in OCD. This spatial specificity suggests the *KNCN/MKNK1-AS1* regulatory axis may be particularly important for the development and function of circuits disrupted in OCD. A recent functional imaging study demonstrated that altered resting-state connectivity specifically involving the putamen, a critical striatal region highlighted by our co-expression analyses, was associated with obsessive-compulsive symptoms in adolescents, particularly under conditions of increased environmental stress.^33^ These converging genetic and neuroimaging results suggest that dysregulated molecular and functional interactions in the striatum may contribute to OCD pathophysiology. Additionally, the enrichment of OCD risk genes among genes co-expressed with this regulatory pair further strengthens the connection to disease pathophysiology.

The functional pathways enriched among *KNCN/MKNK1-AS1* highly co-expressed genes, including synaptic vesicle dynamics, calcium signaling, and cilium organization, align well with emerging models of OCD pathogenesis. Disrupted synaptic transmission and calcium homeostasis have been implicated in OCD through both human genetics and animal models.^34–36^ The unexpected enrichment of cilium-related processes is particularly intriguing, as primary cilia play crucial roles in neuronal development and signaling, though their involvement in OCD has not been previously explored.^37^

Several limitations should be acknowledged. First, while our sample size was sufficient to detect the strongest associations, the deflated genomic inflation factors suggest we may be underpowered to detect additional loci with smaller effect sizes. Larger cohorts will be needed to comprehensively map the contribution of antisense lncRNA variants to OCD risk. Second, our analysis focused on rare variants with evolutionary conservation, potentially missing functionally important variants in less conserved regions. Third, the functional consequences of the identified variants require experimental validation through cellular and animal models. Third, the observed expression correlations between antisense lncRNA and protein-coding gene pairs, as identified from GTEx bulk tissue data, highlight potential regulatory relationships; however, we acknowledge these correlations alone do not definitively establish causative regulatory mechanisms. It remains possible that the strong co-expression signals represent either direct cis-regulatory interactions mediated by antisense transcription or indirect co-regulation through shared transcription factors, enhancer elements, or epigenetic marks. Future functional validation experiments, such as targeted genome editing or antisense RNA knockdown studies, will be critical to confirm direct regulatory causation and delineate the precise molecular mechanisms underpinning these observed expression correlations.

Future studies should prioritize functional characterization of the *KNCN/MKNK1-AS1* variants using patient-derived induced pluripotent stem cells and brain organoids. CRISPR-based editing could be used to model specific variants and assess their effects on gene expression, neuronal development, and circuit function. Additionally, investigating whether antisense lncRNA variants influence OCD symptom dimensions, treatment response, or disease trajectory could enable more personalized therapeutic approaches.

By focusing on antisense lncRNA overlap regions, we identified a novel OCD risk locus. The *KNCN/MKNK1-AS1* association, combined with its striatal-specific expression patterns and co-expression with OCD risk genes, provides new insights into the molecular mechanisms underlying OCD. These findings expand our understanding of the genetic architecture of neuropsychiatric disorders beyond protein-coding variation and highlight antisense lncRNAs as promising targets for next-generation therapeutics. As we move toward precision medicine approaches for psychiatric disorders, targeting specific regulatory RNAs based on individual genetic profiles may offer more effective and personalized treatment strategies for OCD and related conditions.

## Methods

### Study population and data acquisition

The All of Us Research Program, led by the National Institutes of Health (NIH), is a pioneering precision medicine initiative designed to advance health research and medical discoveries by collecting diverse data from at least one million participants across the United States.^38^ In the current All of Us Research Program workspace (controlled tier, version 8), we identified samples diagnosed with OCD, of which 2,764 had available whole-genome sequencing (WGS) data. OCD diagnoses were identified using International Classification of Diseases, 10th Revision (ICD-10) codes (F42.0– F42.9). For case-control matching, we selected these 2,764 OCD cases and 271,481 individuals without mental disorders (ICD-10 F01-F99) from the WGS dataset. Using the R package optmatch, we performed 1:5 case-control matching based on sex and principal components (PCs) 1–5, which were derived from WGS data and provided by the All of Us Research Program.

### Whole-genome sequencing and variant calling

From the whole-genome sequencing (WGS) dataset of 414,830 samples, we retained 2,764 cases and 13,820 matched controls after applying quality control (QC) filters. Individuals were excluded if they had fewer than 2.4 million or more than 5.0 million total variants, over 100,000 variants absent from gnomAD v3.1,^24^ or a heterozygous-to-homozygous variant ratio exceeding 3.3.

We further filtered out low-quality variants based on the following criteria: genotype quality (GQ) ≤ 20, read depth (DP) ≤ 10, allele balance (AB) ≤ 0.2 for heterozygotes, ExcessHet < 54.69, and quality scores (QUAL) < 60 for single nucleotide variants (SNVs) and < 69 for short insertions and deletions (InDels). Variants located in low-complexity regions were also removed. Additional QC steps excluded samples with call rates more than three standard deviations below the mean, individuals with close genetic relatedness (kinship coefficient > 0.1) based on Kinship-based INference for Genome-wide association studies (KING),^39^ and those with sex discrepancies.

After filtering, 2,561 cases and 12,974 controls remained. Further exclusions included homozygous reference calls with < 90% read depth supporting the reference allele or GQ < 25, heterozygous calls with AB < 30%, GQ < 25, or a binomial probability > 1×10⁻ for the observed allele balance centered at 0.5, and homozygous alternate calls with < 90% read depth supporting the alternate allele or GQ < 25. Variants with a call rate ≤ 90% and Hardy-Weinberg equilibrium P < 1×10⁻¹² were also removed.

### Annotation of antisense lncRNAs and variant selection

We utilized the Variant Effect Predictor (VEP) to predict the potential functional effects of the variants.^40^ To filter rare variants, we included those with an allele count of ≤ 5 in both our case-control cohort and the non-psychiatric subset of the gnomAD database v3.1.^24^ Antisense long non-coding RNAs (lncRNAs) were annotated using the Ensembl database.^41^ Transcripts were included if they met the following criteria: ≥200 nucleotides in length, lacking protein-coding potential, and transcribed from the antisense strand of annotated protein-coding genes. Rare variants located within antisense lncRNA transcripts and protein-coding genes were extracted for analysis. Variants in low-complexity regions or those with poor sequencing coverage were excluded from further analysis.

### Statistical analysis

Rare variant burden analyses were conducted using Fisher’s exact tests to compare the cumulative number of rare variants in each antisense lncRNA between OCD cases and controls. Relative risk ratios (RRs) with 95% confidence intervals (CIs) were calculated to quantify the enrichment of rare variants in OCD cases relative to controls using and binomial tests. False discovery rates (FDRs) were controlled using the Benjamini– Hochberg method, with an FDR threshold of <0.1 defining statistically significant associations. Odds ratios (ORs) and 95% CIs were also computed to further quantify associations.

For gene constraint analysis, cognate sense genes were stratified into deciles based on their loss-of-function observed/expected upper bound fraction (LOEUF) scores (range: 0–2, where lower scores indicate stronger selective constraint)^24^ and AlphaMissense scores (range: 0–1, where higher scores reflect stronger selective pressure against missense variants).^42^ Enrichment analyses were performed within each decile to identify differential burden patterns among OCD-associated antisense lncRNAs using binomial tests.

Gene-based association testing was performed using the optimal unified sequence kernel association test (SKAT-O) implemented in the SKAT R package. SKAT-O combines the burden test and SKAT through an optimal linear combination, providing robust power across different genetic architectures. The test adaptively selects the optimal combination parameter that maximizes statistical power with the covariate matrix including sex and PC1-PC5. Only genes harboring at least two rare variants (allele count ≤ 5) were included in the analysis. Three test statistics were computed for each gene: (1) SKAT p-value, optimal for scenarios where variants have different directions of effect; (2) Burden test p-value, optimal when variants have the same direction of effect; and (3) SKAT-O p-value, which adaptively combines both tests.

To account for multiple testing across genes, we applied two correction methods. The Bonferroni correction was calculated as α/n, where α = 0.05 and n represents the number of genes tested. Additionally, the false discovery rate (FDR) was controlled using the Benjamini-Hochberg procedure. Genes with FDR <0.05 were considered statistically significant.

We employed both Fisher’s exact test and SKAT-O for region-based rare variant association testing across antisense lncRNA-protein coding overlap regions. Fisher’s exact test compares the proportion of individuals carrying at least one rare variant within each overlap region between cases and controls, assuming all variants contribute similarly to disease risk. SKAT-O combines burden and variance-component approaches at the gene level, providing robust power across different genetic architectures—whether all rare variants in a region increase risk (burden model optimal) or when a region contains both risk and protective variants (variance component model optimal). When interpreting discordant results, SKAT-O significance with non-significant Fisher’s exact test suggests genetic heterogeneity within the region, while Fisher’s significance alone indicates consistent directional effects across variants.

### Transcriptomic analyses

To investigate the correlation between *KNCN* and *MKNK1-AS1* gene expression across multiple brain tissues, RNA-seq data from the Genotype-Tissue Expression (GTEx) v10 dataset were utilized. For each of the 13 brain tissues, gene expression data were obtained in transcripts per million (TPM) format. Pearson correlation coefficients were calculated for each tissue to quantify the relationship between *KNCN* and *MKNK1-AS1* expression using “cor.test” function in R. For the correlation analysis, expression data for each tissue were log-transformed (log2(TPM + 1)) to normalize distribution. Statistical significance of the correlations was determined by performing Pearson’s correlation tests.

To identify genes demonstrating strong correlation with both *KNCN* and *MKNK1-AS1* expression across four brain tissues (the nucleus accumbens, hypothalamus, putamen, and caudate) with Pearson r > 0.7 between *KNCN* and *MKNK1-AS1* expression, we conducted a comprehensive correlation analysis. We calculated Pearson correlation coefficients between the expression of each gene and both *KNCN* and *MKNK1-AS1* individually within each tissue. Genes, with median TPM ≥ 0.1, exhibiting strong correlations (|Pearson r| > 0.7) with both *KNCN* and *MKNK1-AS1* were identified and saved separately per tissue.

We then performed Gene Ontology (GO) enrichment analysis separately for each tissue using its respective set of high-correlation genes. This tissue-specific approach allowed us to identify biological processes that may be regulated by the *KNCN/MKNK1-AS1* axis in a region-dependent manner. Enriched GO terms were filtered for biological processes with FDR < 0.05 and compared across tissues to identify both shared and tissue-specific pathways.

### Comparison with OCD risk genes identified in GWAS

The current GWAS identified 207 genes associated with OCD risk.^6^ To evaluate whether OCD-associated genes were significantly enriched among genes showing high correlation with both *KNCN* and *MKNK1-AS1* expression in the four brain tissues (the nucleus accumbens, hypothalamus, putamen, and caudate), we performed tissue-specific enrichment analyses using Fisher’s exact test. For each tissue, we first identified the set of genes, with median TPM ≥ 0.1, demonstrating strong correlation (|Pearson r| > 0.7) with both *KNCN* and *MKNK1-AS1* expressions as described above.

We constructed a 2 × 2 contingency table for each tissue, classifying genes according to two criteria: (1) whether they were OCD-associated genes and (2) whether they showed high correlation with both *KNCN* and *MKNK1-AS1*. The contingency table comprised four categories: (a) OCD-associated genes present in the high-correlation set; (b) non-OCD genes present in the high-correlation set; (c) OCD-associated genes not present in the high-correlation set but expressed in the tissue; and (d) non-OCD genes expressed in the tissue but not in the high-correlation set.

Using Fisher’s exact test, we assessed whether the overlap between the OCD gene set and the high-correlation gene set for each tissue was greater than expected by chance. We corrected for multiple testing across the four tissues using the Benjamini-Hochberg procedure to control the FDR. Enrichment results with an FDR-adjusted *P* value < 0.05 were considered statistically significant.

## Data Availability

All of Us controlled tier data is available for registered institutions and cohort selection is detailed in the workbench titled "Detecting the prevalence of ultra-rare gene mutations." Due to stringent privacy protections and data sharing restrictions enforced by the All of Us Research Program, individual-level variant data cannot be shared publicly. Researchers must adhere to specific guidelines to protect participant confidentiality, which includes not reporting or disseminating any data that could allow the re-identification of participants, particularly those involving participant counts between 1 and 20 without employing approved data obscuration methods.

## Acknowledgements

This study was supported by a grant from Beatrice and Samuel A. Seaver Foundation (BM).

The All of Us Research Program is supported by the National Institutes of Health, Office of the Director: Regional Medical Centers: 1 OT2 OD026549; 1 OT2 OD026554; 1 OT2 OD026557; 1 OT2 OD026556; 1 OT2 OD026550; 1 OT2 OD 026552; 1 OT2 OD026553; 1 OT2 OD026548; 1 OT2 OD026551; 1 OT2 OD026555; IAA #: AOD 16037; Federally Qualified Health Centers: HHSN 263201600085U; Data and Research Center: 5 U2C OD023196; Biobank: 1 U24 OD023121; The Participant Center: U24 OD023176; Participant Technology Systems Center: 1 U24 OD023163; Communications and Engagement: 3 OT2 OD023205; 3 OT2 OD023206; and Community Partners: 1 OT2 OD025277; 3 OT2 OD025315; 1 OT2 OD025337; 1 OT2 OD025276. In addition, the All of Us Research Program would not be possible without the partnership of its participants.

## Author contributions

Study concept and design: SJ, BM

Acquisition, analysis, or interpretation of data: MC, SJ, BM

Drafting of the manuscript: All authors

Critical revision of the manuscript for important intellectual content: All authors

Statistical analysis: MC, SJ, BM

Obtained funding: BM

Study supervision: BM

## Code availability

Code and resources used in this study is available on GitHub at (https://github.com/MahjaniLab/OCD_WGS). For any further inquiries or requests for code not available in the repository, please contact the corresponding author.

