## Supplemental Figures for "Rare Variants in Antisense lncRNA-Protein Coding Gene Overlap Regions Contribute to Obsessive-Compulsive Disorder"

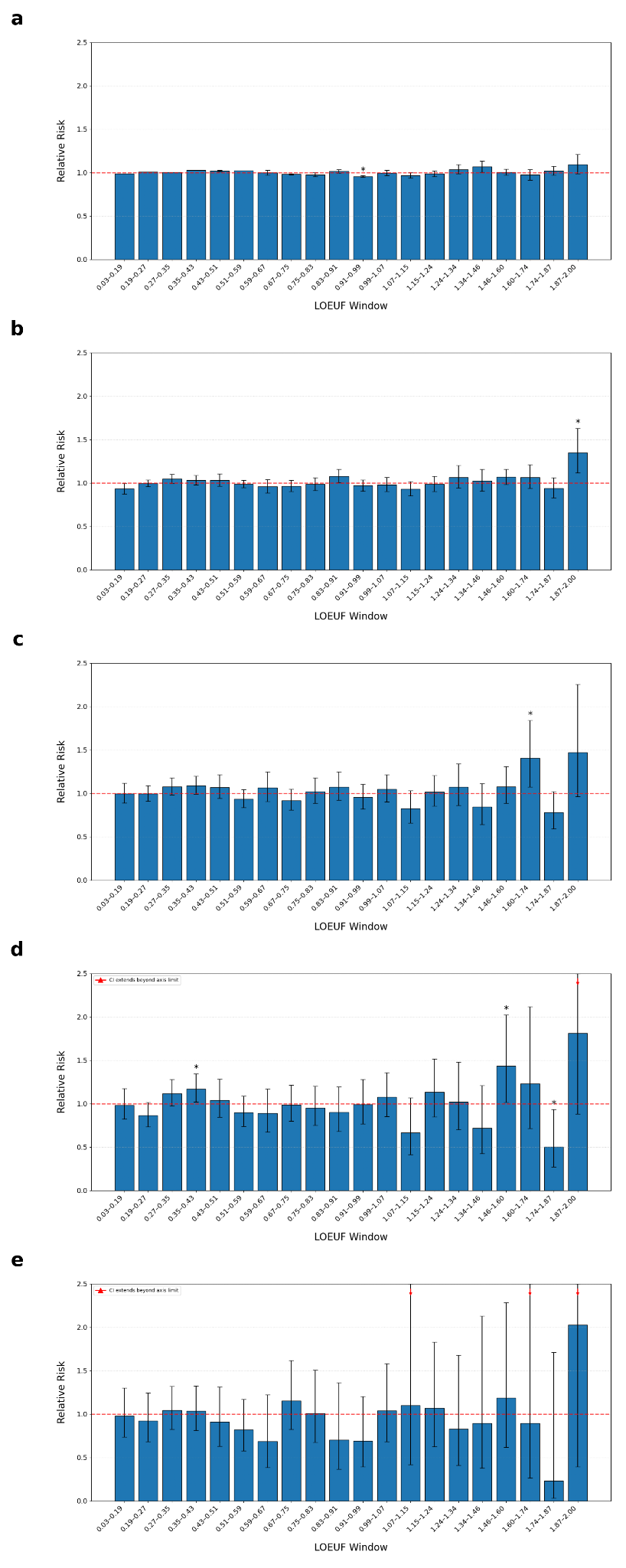

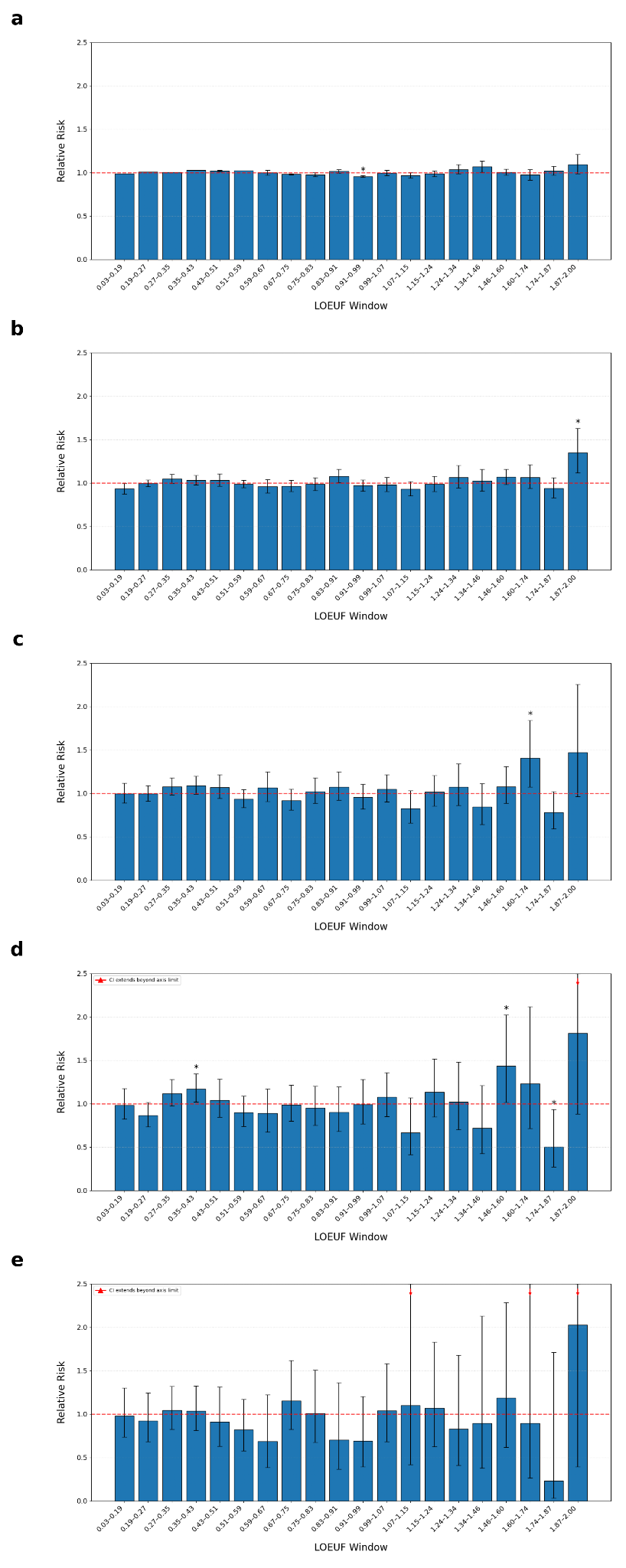

**Supplementary Figure 1. Rare variant burden in antisense lncRNA overlap regions stratified by gene constraint levels and conservation scores.** Analysis of rare variant burden in antisense lncRNA overlap regions across genes stratified by constraint levels. Genes were divided into 20 quantiles based on their LOEUF (Loss-of-function Observed/Expected Upper bound Fraction) scores, where lower LOEUF values indicate greater constraint against loss-of-function variation. (a) Relative risk of rare variants in antisense lncRNA overlap regions across all LOEUF quantiles, showing no conservation filter applied (all variants included regardless of GERP++ score). (b) Relative risk analysis with conservation filter requiring GERP++ score > 0. (c) Relative risk analysis with conservation filter requiring GERP++ score > 1. (d) Relative risk analysis with conservation filter requiring GERP++ score > 2, showing emerging enrichment in the least constrained genes (20th quantile). (e) Relative risk analysis with the most stringent conservation filter (GERP++ score > 3).

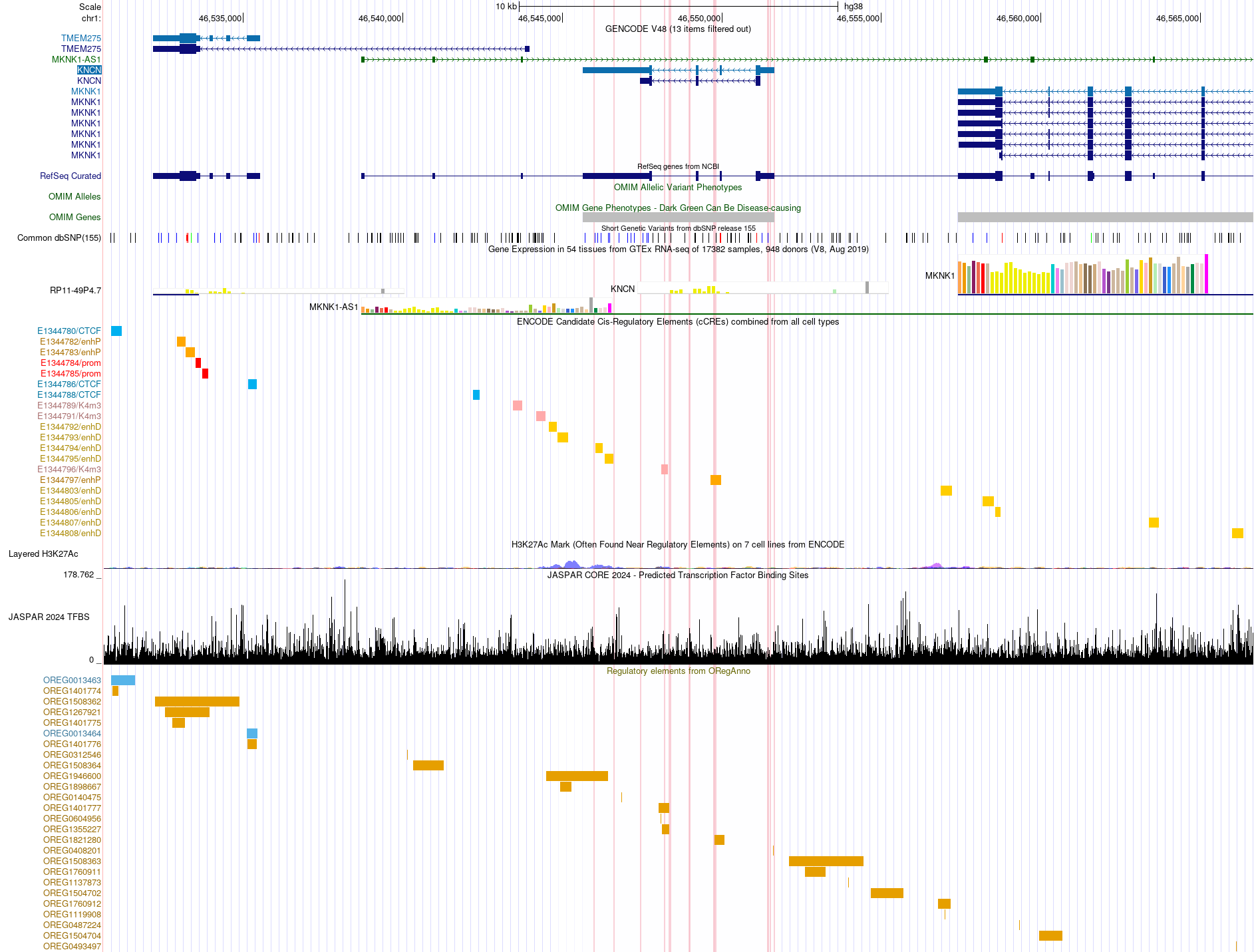

**Supplementary Figure 2. Integrated genomic landscape of the *KNCN/MKNK1-AS1* locus revealing regulatory architecture and transcription factor binding sites.** Multi-track visualization displaying the genomic region chr1:46,530,623-46,566,664 (hg38/GRCh38). GENECODE v48 and RefSeq gene annotations show *KNCN*, *MKNK1-AS1*, *TMEM275* and *MKNK1* gene structures with exons (thick bars) and introns (thin lines). Common genetic variants from dbSNP build 155 are displayed along with additional transcript annotations. ENCODE candidate cis-regulatory elements (cCREs) are integrated from multiple cell types, with each colored block representing a distinct regulatory element class. H3K27ac ChIP-seq signal (yellow/orange) from seven ENCODE cell lines indicates active enhancer regions. JASPAR 2024 predicted transcription factor binding sites are shown with binding affinity scores (black histogram) and individual TF motifs color-coded by factor identity. Vertical pink lines denote regions including rare conserved variants identified in this study.

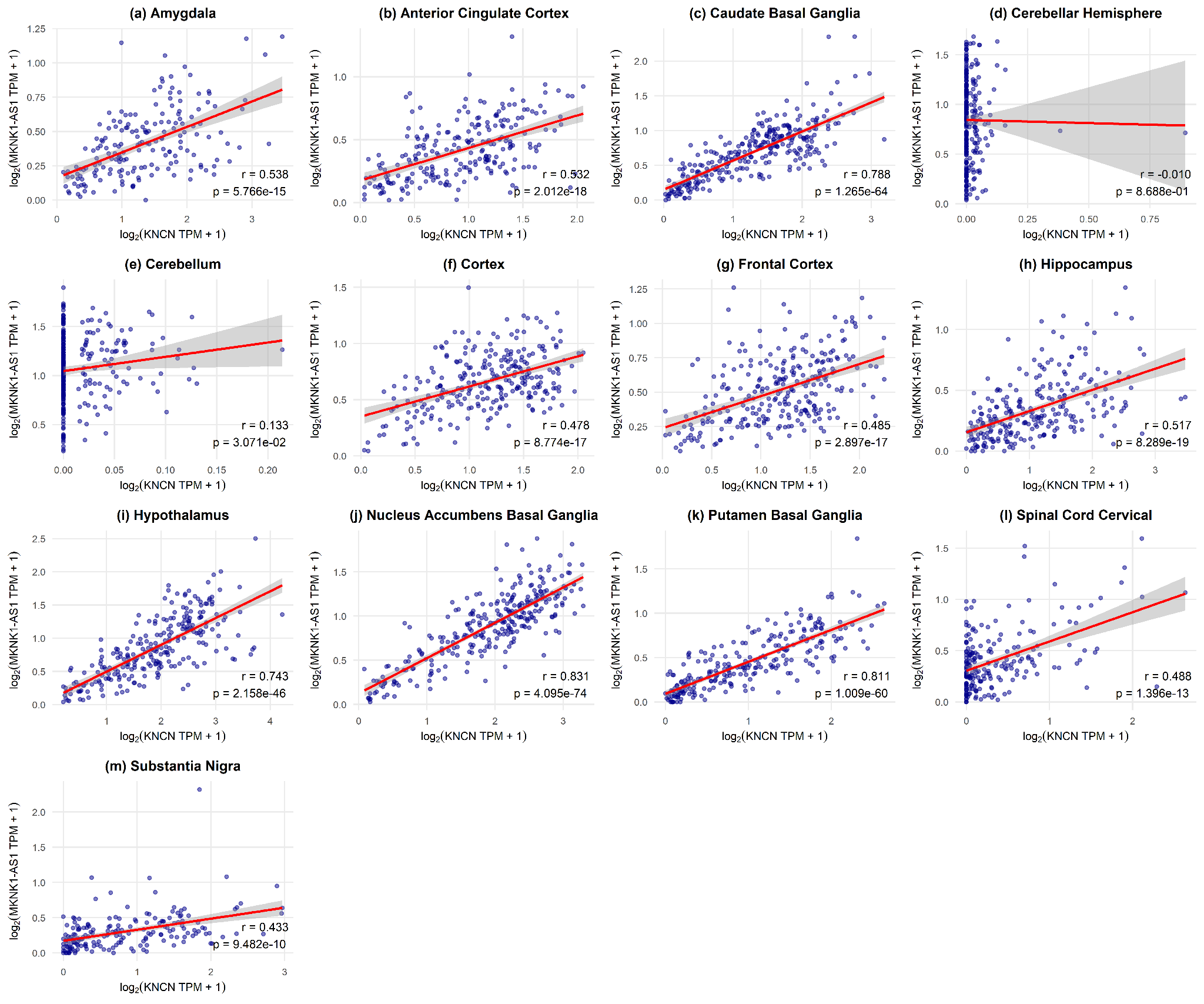

**Supplementary Figure 3. Correlation between KNCN and MKNK1-AS1 expression across 13 human brain regions.** Scatter plots showing the relationship between log2-transformed expression values (TPM + 1) of KNCN (x-axis) and MKNK1-AS1 (y-axis) in (a) amygdala, (b) anterior cingulate cortex, (c) caudate basal ganglia, (d) cerebellar hemisphere, (e) cerebellum, (f) cortex, (g) frontal cortex, (h) hippocampus, (i) hypothalamus, (j) nucleus accumbens basal ganglia, (k) putamen basal ganglia, (l) spinal cord cervical c-1, and (m) substantia nigra. Each point represents an individual sample from the GTEx v10 database. Red lines indicate linear regression fits with 95% confidence intervals shown in gray shading. Pearson correlation coefficients (r) and p-values are displayed for each brain region. Panels are arranged alphabetically by brain region name.
